# COVID-19 risk assessment for the Tokyo Olympic Games

**DOI:** 10.1101/2021.04.14.21255261

**Authors:** Wenhui Zhu, Jie Feng, Cheng Li, Huimin Wang, Yang Zhong, Xingyu Zhang, Tao Zhang

## Abstract

**Objectives:** To explore effective prevention and control measures for Coronavirus Disease 2019 (COVID-19) in large international events through simulations of different interventions according to risk assessment.

**Methods:** We used random model to calculate the number of initial infected patients. And Poisson distribution was used to determine the number of initial infected patients based on the number of countries involved. Further, to simulate the COVID-19 transmission, the susceptible-exposed-symptomatic-asymptomatic-recovered-hospitalized (SEIARH) model was established based on susceptible-exposed-infectious-recovered (SEIR) mathematical model of epidemic diseases. According to risk assessment indicators produced by different scenarios of the simulated interventions, the risk of COVID-19 transmission in Tokyo Olympic Games was assessed.

**Results:** The large-scale vaccination will effectively control the spread of COVID-19. If the effective rate of vaccine is 100%, and the vaccination rate of athletes reaches 80%, an epidemic prevention barrier can be established.

**Conclusions:** The current COVID-19 prevention measures proposed by the Japan Olympic Committee were needed to be enhanced. For the vaccination intervention had the best control effect, a mass vaccination serves was an effective way to control COVID-19.

## Background

The 32nd Summer Olympic Games (Games of the XXII Olympiad) will be held from July 23 to August 8, 2021 in Tokyo, Japan [1]. However, the Olympics Games is easily exposed by the Coronavirus Disease 2019 (COVID-19). Till March 30, 2021, the outbreak of COVID-19 has spread to more than 200 countries [2]. The global epidemic situation remains grim, with more than 12.3 million global cumulative reported cases and more than 2.8 million deaths [3], while the number of infected individuals is still growing rapidly. Consequently, numerous major events and activities around the world have been cancelled or postponed, and preparations for all parts of upcoming sport events are greatly challenged. Under such circumstances, about 11,000 athletes from approximately 206 countries and regions are arriving in Tokyo for the 32nd Summer Olympic during the summer of 2021 [4], along with coaches, referees, and associated International Sport Organization officials, which will definitely increase the risk of infectious disease outbreaks and transmissions. Although the International Olympic Committee (IOC) has made it clear that overseas audiences will not be allowed to enter Japan to view the Tokyo Olympic Games [5], it is far from enough to ensure the safety of Olympic Games in terms of COVID-19. Therefore, it is urgently necessary to commence an evaluation for the risk of COVID-19 under different prevention measures.

So far the COVID-19 prevention measures proposed by the Japan Olympic Committee (JOC) includes the following aspects [6]:

1. “Keep a minimum of two metres’ distance from athletes at all times. Keep a minimum of one metres’ distance from others.
2. Be ready to take a COVID-19 test at the airport, depending on the country you traveled from and where you have been in the last 14 days. Be ready to show immigration authorities: Evidence of your negative COVID-19 test taken within 72 hours of your departure.
3. During your stay in Japan, you will be expected to limit your activities to what is required to carry out your role.”

Although there was no specific plan for vaccination by the IOC and JOC, Thomas Bach, the president of IOC, said that if a vaccine becomes available in time for the 23 July-8 August Games in 2021, the IOC would foot the bill [7]. However, since vaccination is voluntary for atheletes, different vaccine coverage may produce different effects on the outbreak of the Olympic Village. In the light that how it will be has not been studied so far, we used different vaccine coverage to simulate the transmission of COVID-19 in this study.

In this paper, to assess the risk of COVID-19 during the Tokyo Olympic Games, a simulation study based on the transmission dynamic model was carried out. Firstly, we collected the number of athletes from different countries participating in the Tokyo Olympic Games, the current COVID-19 infection probability of each country, and the transmission parameters of the COVID-19 model. Secondly, utilizing the initial number of asymptomatic infections, the number of contacts and other aspects, we established the susceptible-exposed-symptomatic-asymptomatic-recovered-hospitalized (SEIARH) model. Thirdly, in order to carry out risk assessments, the secondary infectors at peak hour and peak hour of onset were calculated. Through realizing a comparison of the expected risks of COVID-19 under different prevention strategies, this study provided quantitative reference evidence regarding the formulation of COVID-19 prevention and control programmes for the Tokyo Olympic Games.

## Methods

The methods of this study consisted of three parts. Firstly, to determine the number of initial infected patients based on the number of countries coming to Japan the Poisson distribution was used. Secondly, based on COVID-19 transmission mechanism, SEIARH model was established to simulate COVID-19 transmission. Thirdly, according to the results of SEIARH model, the risk assessment indicators (secondary infectors at peak hour, peak hour of onset / d) were calculated.

### Data collection and preparation

National infection probability *π*_*i*_ (*i* = 1, 2,…, *n*) was calculated according to public data[3]:

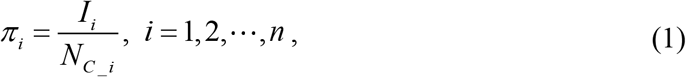

where *I*_*i*_ is the number of infections in country *i*; *N*_*C* _ *i*_ is the total population number of country *i*.

The Olympic Games are held every four years, little change existed in the total number of the adjacent two sessions [8]. Besides, the number of participants was not available before the announcement of IOC, so we used the number of participants announced in the 2016 Rio Olympic Games as a reference [9]. Further, the initial asymptomatic infected population was assumed to 10 for the previous imported asymptomatic infected cases did not exceed 10 at one time.

For the parameters of SEIARH model, we refered to the published COVID-19 classic retrospective study.

### Determining the number of initial infected patients

Assuming the presence of COVID-19 infectors at entry who are not identified by entry quarantine or health check-up. Here we combined the probability of infection to calculate the number of overseas import infectors. Furthermore, the initial infected patients referred to unidentified infectors among athletes, coaches, referees, officials and others who enter Japan for the Tokyo Olympics Games, since the number of participants and the epidemics of COVID-19 vary across different countries.

The number of infected patients assumed from different regions *i*(*i* = 1, 2,…, *n*) is *X*_*i*_. It can be approximately seen that the Poisson distribution of compliance parameter *λ*_*i*_= *N*_*i*_*π* _*i*_, which is in the form of:

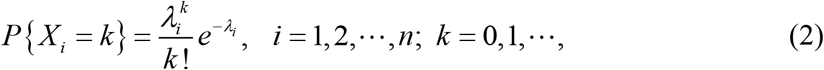

where *N*_*i*_ is the number of athletes, coaches, referees, and officials from different regions; *π*_*i*_ is the infection probability of immigrants from different regions. The infected input probability *P*_*i* _ *imported*_ of person from the region *i* was:

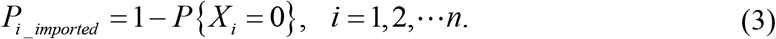

Furthermore, the total number of initial infected patients could be represented as:

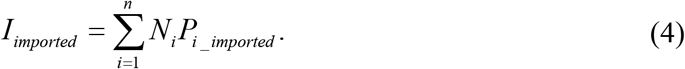

### Simulating the transmission of COVID-19 during the Tokyo Olympic Games

Based on the transmission mechanism, individual epidemiological status and prevention and control measures of COVID-19 infection, we established a SEIARH warehouse model [10]-[11]. The population was partitioned into subpopulations as: susceptible (S), exposed (E), symptomatic infected (I), asymptomatic infected (A), hospitalized (H) and recovered (R). The transmission process is shown in **Fig. 1**.

**Fig. 1.**
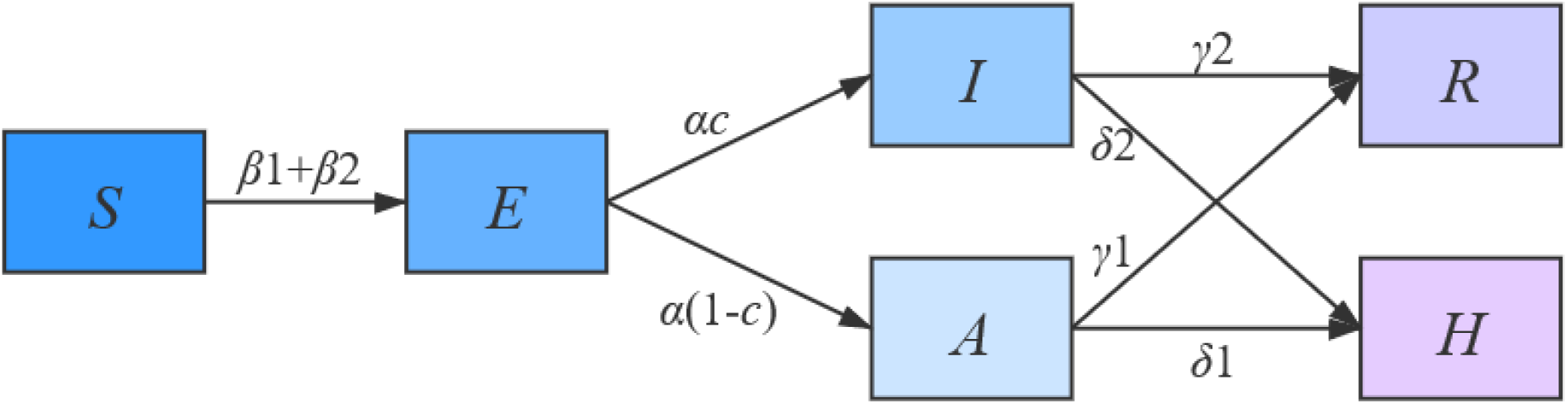
Flow diagram of SEIARH model.

The discrete-time stochastic compartment model for COVID-19 infection was constructed as:

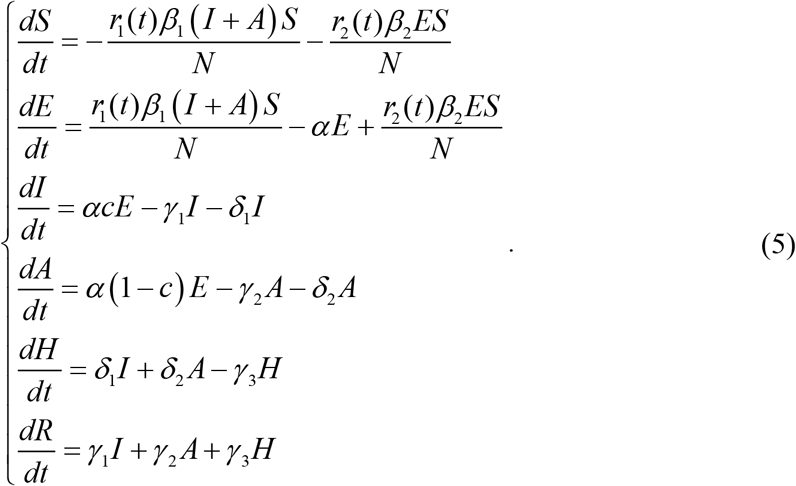

The parameters were defined in **Table 1**. As the Japan authority has proposed that “During your stay in Japan, you will be expected to limit your activities to what is required to carry out your role.” [6]. Therefore, the range of activities of athletes outside the Olympic village was not considered, that is the total population number *N*=11,000.

**Table 1.** Parameter definition and estimation

| Parameter | Definition | Value | Source |
| --- | --- | --- | --- |
| $N$ | The total population number | $1.1 \times 10^4$ | [4] |
| $\beta_1$ | Probability of transmission from infected individuals to susceptible individuals | 0.15747 | [12] |
| $\beta_2$ | Probability of transmission from exposed individuals to susceptible individuals | 0.78735 | [12] |
| $r_1(t)$ | Average number of infected class contact with susceptible class | 2.20 | [13] |
| $r_2(t)$ | Average number of infected class contact with exposed class | 2.22 | [14] |
| $\alpha$ | Probability of susceptible individuals becoming infected individuals | 1/5.2 | [15] |
| $\delta_1$ | Transition rate of symptomatic infected individuals to the quarantined infected class | 0.6 | [16] |
| $\delta_2$ | Transition rate of asymptomatic infected individuals to the quarantined infected class | 0.4 | [16] |
| $\gamma_1$ | Recovery rate of symptomatic infected individuals | 0.1 | [17] |
| $\gamma_2$ | Recovery rate of asymptomatic infected individuals | 0.1 | [17] |
| $\gamma_3$ | Recovery rate of hospitalized individuals | 0.15 | [18] |
| $c$ | Probability of infected class have symptoms | 0.4 | [19] |
| $S(0)$ | Initial susceptible population | $1.1 \times 10^4$ | [4] |
| $E(0)$ | Initial exposed population | 0 | |
| $A(0)$ | Initial asymptomatic infected population | 10 | |
| $I(0)$ | Initial symptomatic infected population | 0 | Assumed. |
| $R(0)$ | Initial recovered population | 0 | |
| $H(0)$ | Initial quarantined infected population | 0 | |

\* Note: This study mainly considered the effect of different measures on the prevention and control of the Tokyo Olympic Games in the case of missed detection of asymptomatic infectors, so the initial asymptomatic infectors were 10, and the remaining initial values was 0.

Considering the current regulations on entry of relevant personnel during the games, nucleic acid test results within 72 hours should be submitted before entry, and the number of contacts should be strictly controlled after entry [20]-[21], we defined the number of susceptible individuals exposed by infected (*r*_1_ (*t*)) and exposed (*r*_2_ (*t*)) as a piecewise functions of time *t*, where *t* is the number of days after entry. The function forms were given as follows:

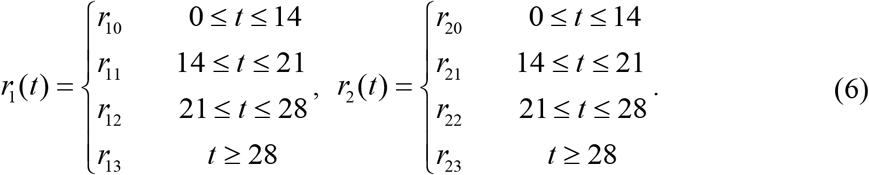

All the above statistical analyses were performed in Matlab R 2016a.

### Assessing the risks of COIVD-19 under different prevention measures

Based on the results of SEIARH model, we calculated the risk assessment index of COIVD-19: secondary infectors at peak hour and peak hour of onset. Secondary infectors at peak hour referred to the maximum number of infected persons after entry into Japan; Hour of onset is the time *t* corresponding to the maximum number of infections. The specific calculation formula was as follows:

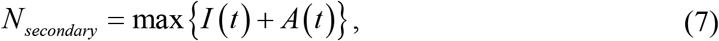

where *N*_*secondary*_ is the number of secondary infectors at peak hour; *I* (*t*) is the number of symptomatic infected population; *A*(*t*) is the number of asymptomatic infected population.

## Results

### Transmission simulation without intervention

Based on ignoring the activities of athletes outside the Olympic Village, we used the parameters of the SEIARH model in **Table 1** to simulate the transmission of the Olympic Village in Japan without intervention, as shown in **Fig. 2**. The number of new secondary infectors reached the peak on the 13th day, which was 1,698; the cumulative infectors were in the S-shaped curve.

**Fig. 2.**
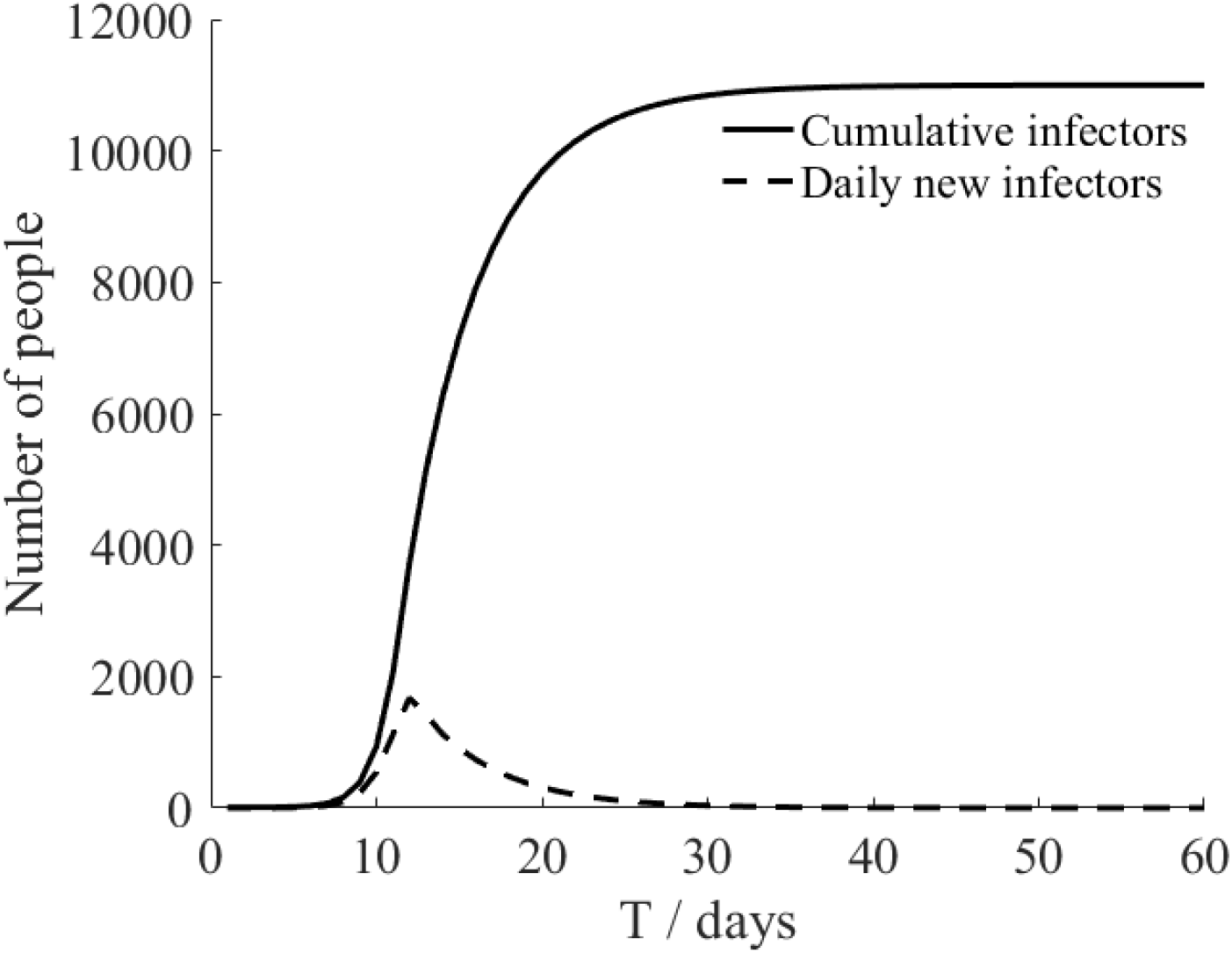
Transmission without intervention.

### Transmission simulation under current interventions of Japan Olympic Committee

According to the current prevention and control measures of JOC, the parameters of average number of infected class contact with susceptible class (*r*_1_), average number of infected class contact with exposed class (*r*_2_), initial asymptomatic infected population (*A*(0)) were adjusted, and the specific settings were shown in **Table 2**.

**Table 2.**
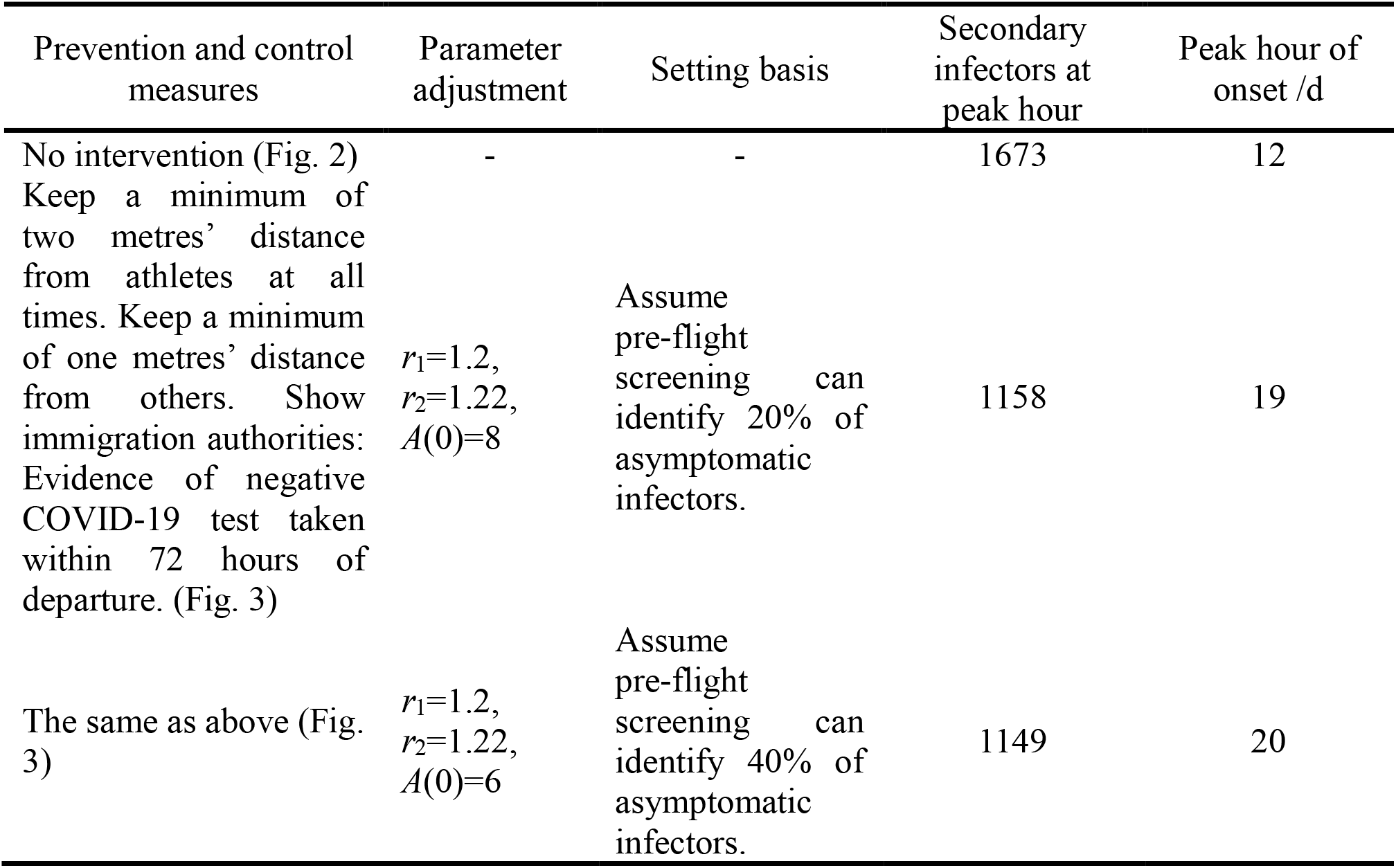
Parameter adjustment and transmission simulation

| Prevention and control measures | Parameter adjustment | Setting basis | Secondary infectors at peak hour | Peak hour of onset /d |
| --- | --- | --- | --- | --- |
| No intervention (Fig. 2) | - | - | 1673 | 12 |
| Keep a minimum of two metres' distance from athletes at all times. Keep a minimum of one metres' distance from others. Show immigration authorities: Evidence of negative COVID-19 test taken within 72 hours of departure. (Fig. 3) | $r_1=1.2$,<br>$r_2=1.22$,<br>$A(0)=8$ | Assume pre-flight screening can identify 20% of asymptomatic infectors. | 1158 | 19 |
| The same as above (Fig. 3) | $r_1=1.2$,<br>$r_2=1.22$,<br>$A(0)=6$ | Assume pre-flight screening can identify 40% of asymptomatic infectors. | 1149 | 20 |

As can be seen from **Fig. 3**, based on the measures currently proposed by JOC, it was assumed that the screening before take-off can reduce 20% and 40% of asymptomatic infectors respectively, but the prevention and control effect was not very ideal. The total number of secondary infectors still reached the level of no intervention measures, but the duration increased; the peak value of daily new secondary infectors decreased, reaching 1,158 on the 19th day and 1,149 on the 20th day, respectively. Through the simulation of the current prevention and control measures, it can be found that it was not enough to rely solely on the existing measures. Other measures (such as vaccination) must be supplemented to effectively reduce the risk of epidemic during the Tokyo Olympic Games. We would discuss it further in next section.

**Fig. 3.**
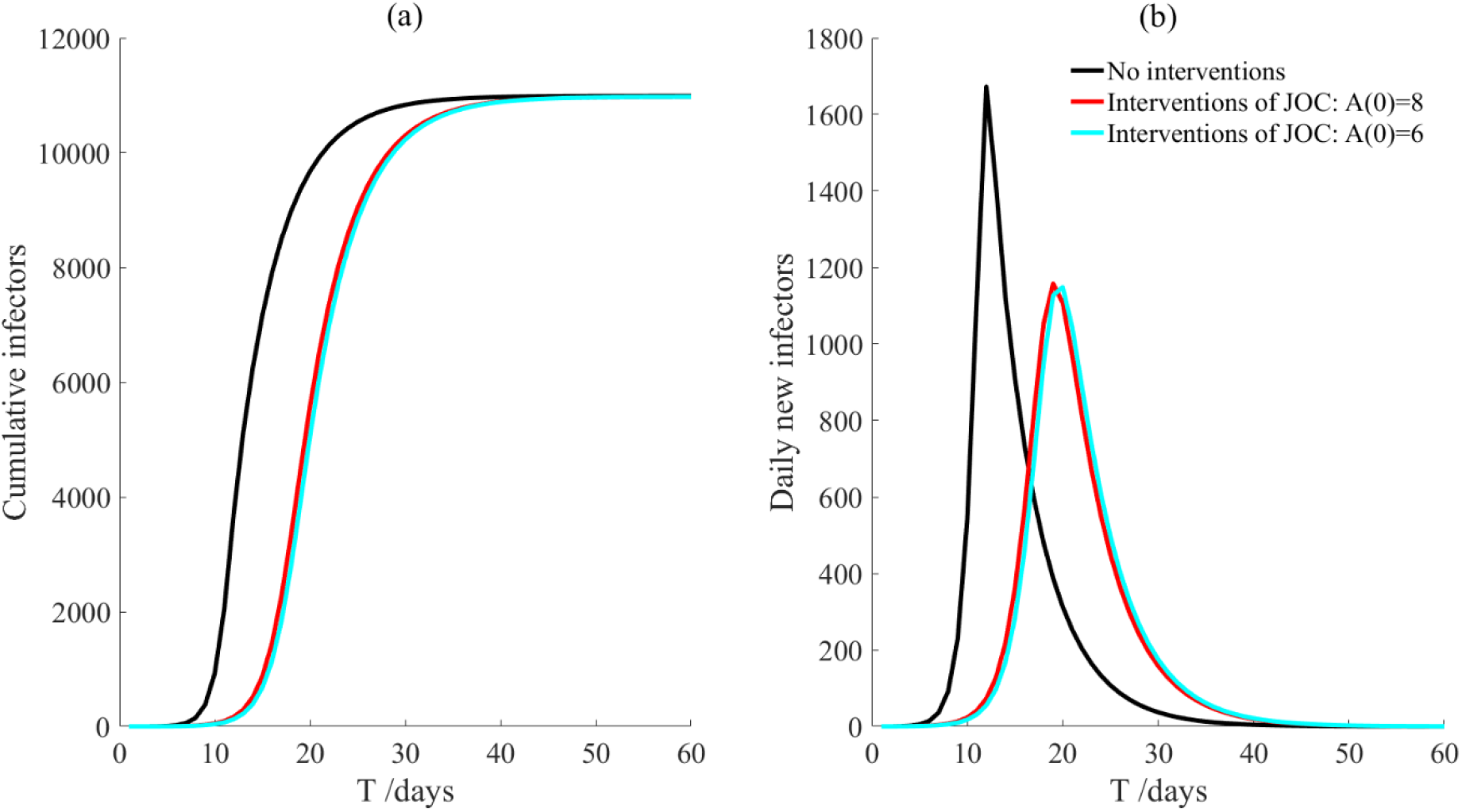
The transmission after JOC interventions.

### Transmission simulation under other interventions

According to the simulation of intervention measures set in the above sections, we can find that the current prevention and control effect was not ideal. But for various infectious diseases, vaccine is the most effective way to eradicate the transmission of infectious diseases. And Thomas Bach, President of the International Olympic Committee, said that if a vaccine becomes available in time for the 23 July-8 August Games in 2021, the IOC would foot the bill [7]. Therefore, based on the intervention measures proposed by JOC, we considered the transmission of athletes after vaccination. The transmission simulation was carried out by adding prevention and control scenarios such as “vaccinated but not quarantined”, “vaccination and 7-day compulsory quarantine”, “vaccination and 14-day compulsory quarantine”. The specific parameter adjustments were shown in **Table 3**.

**Table 3.**
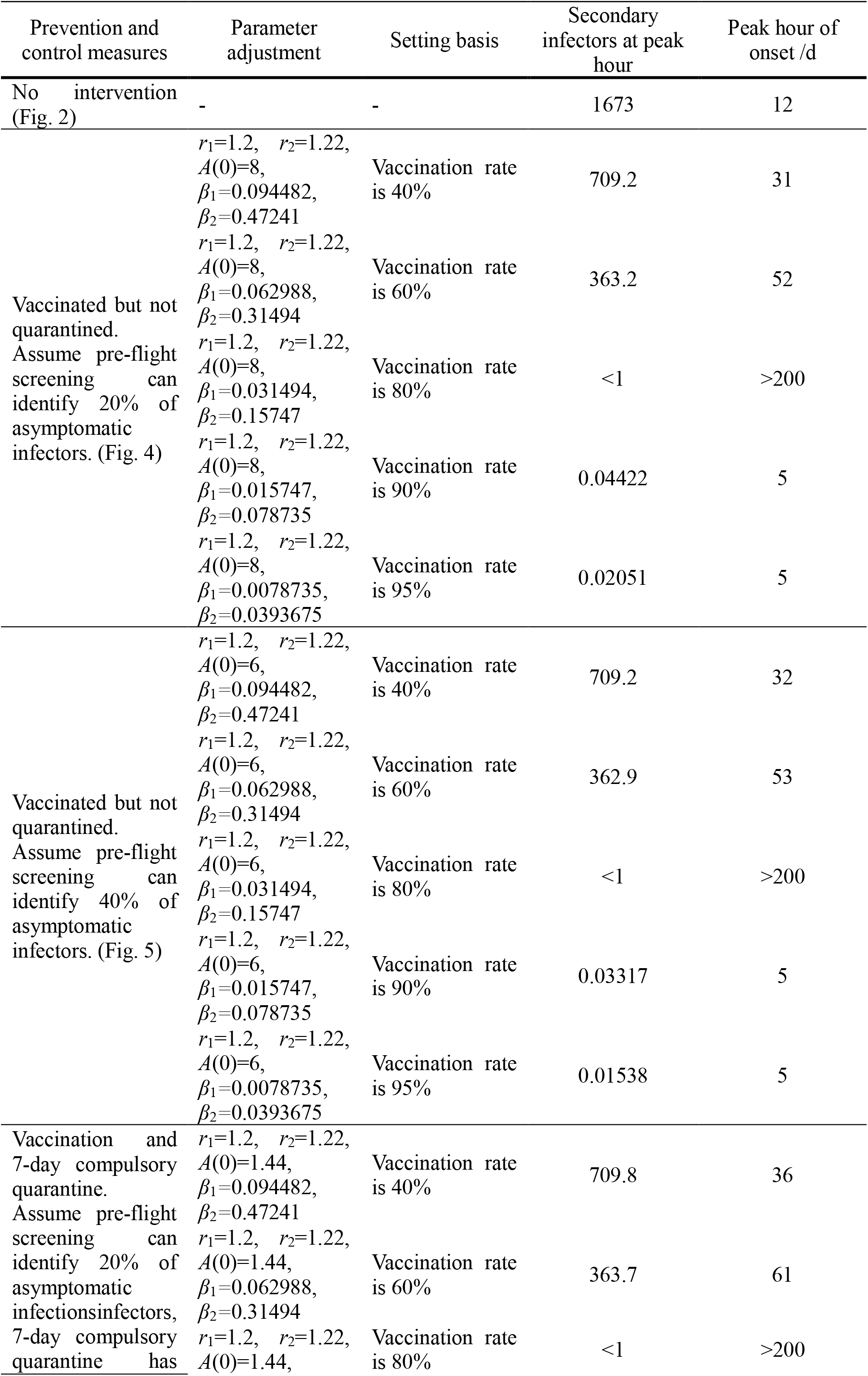
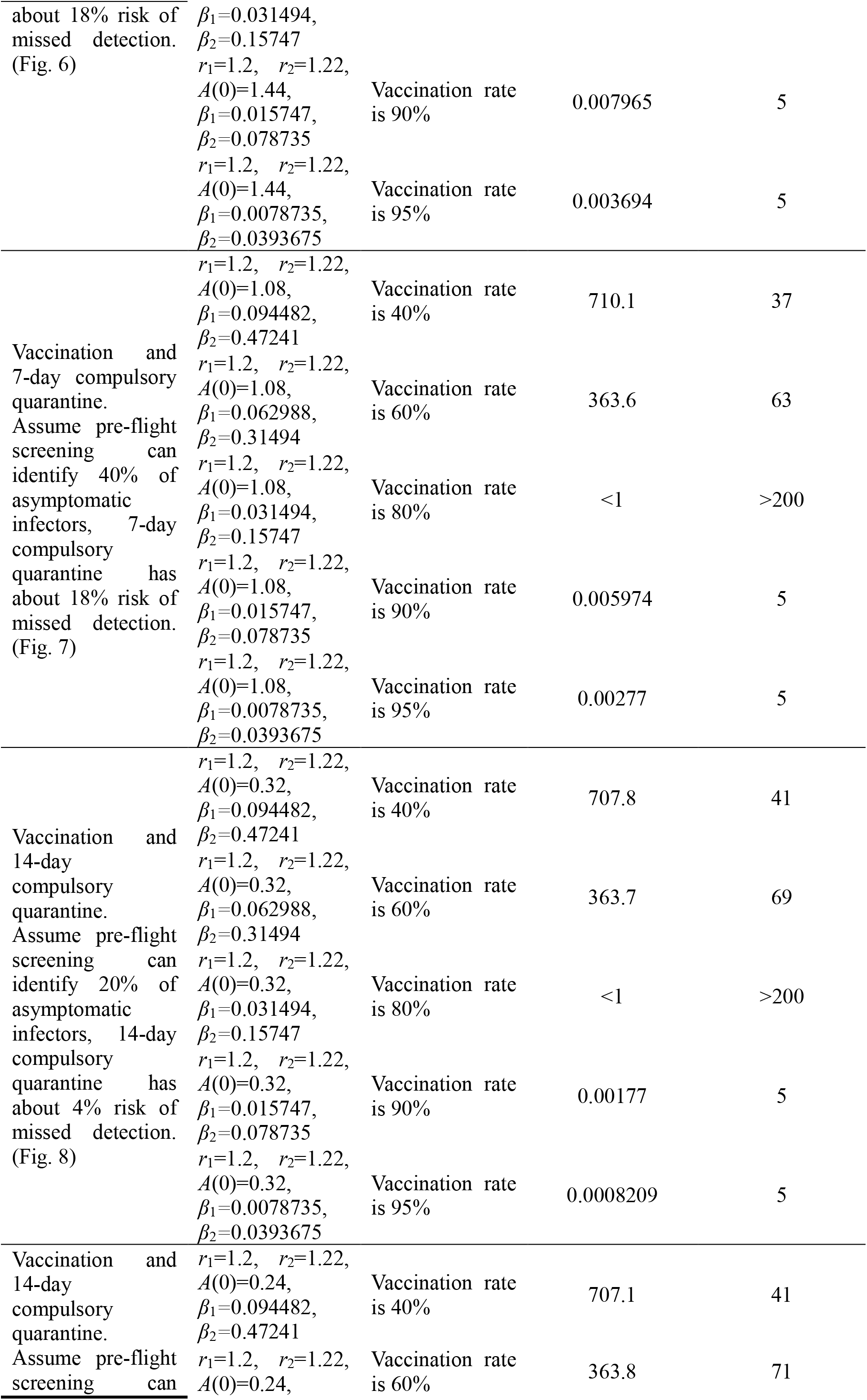
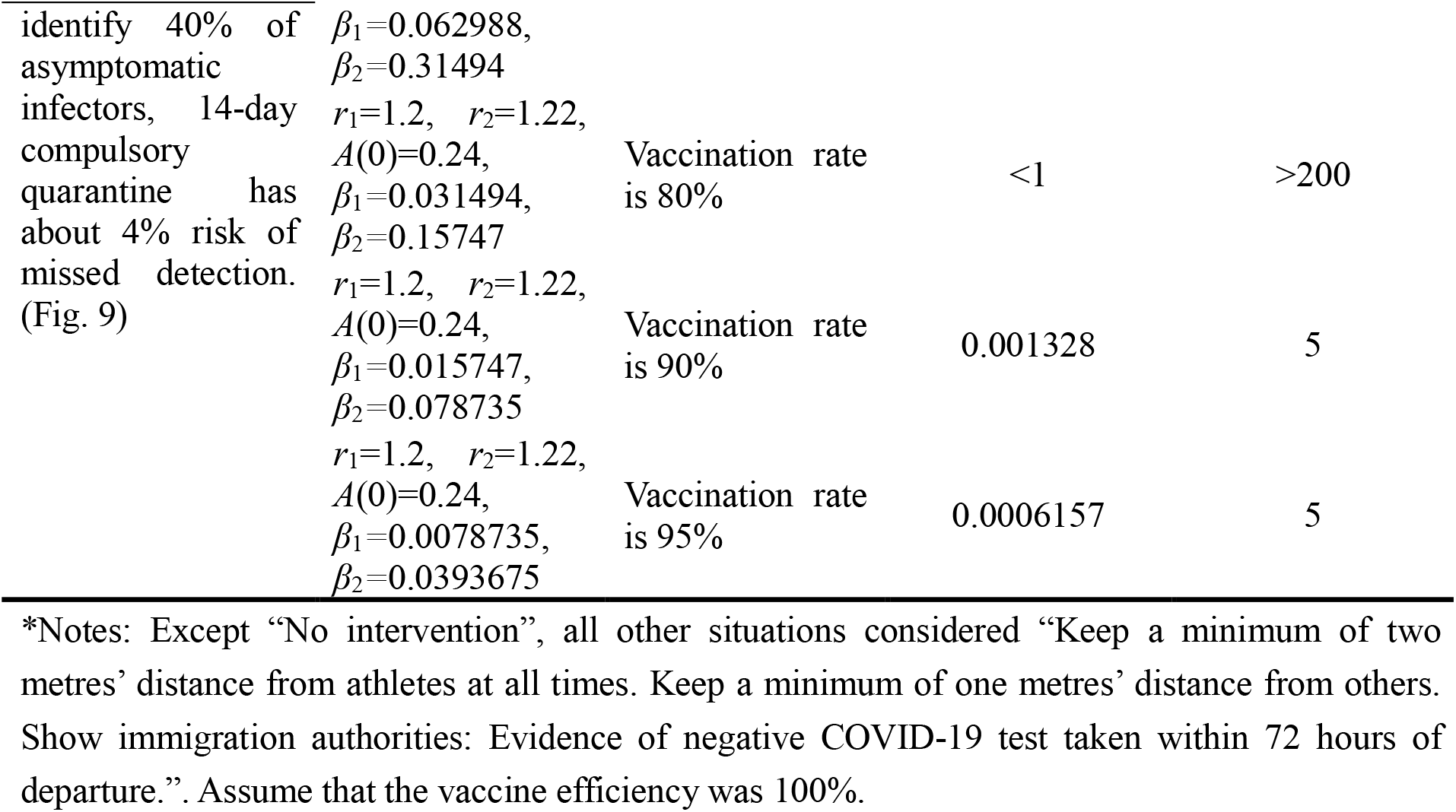
Parameter adjustment and transmission after vaccination

It can be found from **Fig. 4** to **Fig. 9** that under the assumption the vaccine efficiency was 100%. Whether isolated or not, screening before take-off can identify 20% or 40% of asymptomatic infectors, which significantly reduced the number of secondary infectors at peak hour and delayed the peak hour of onset compared with no interventions and current interventions by the JOC. In the comparison of different vaccination rates of various measures, it can be found that when the vaccination rate of athletes reached 80%, the number of secondary infectors at peak hour has decreased significantly. It showed that the effect of the vaccine was obvious, and the extensive vaccination would be useful for the prevention and control of COVID-19 in Tokyo Olympic Games.

**Fig. 4.**
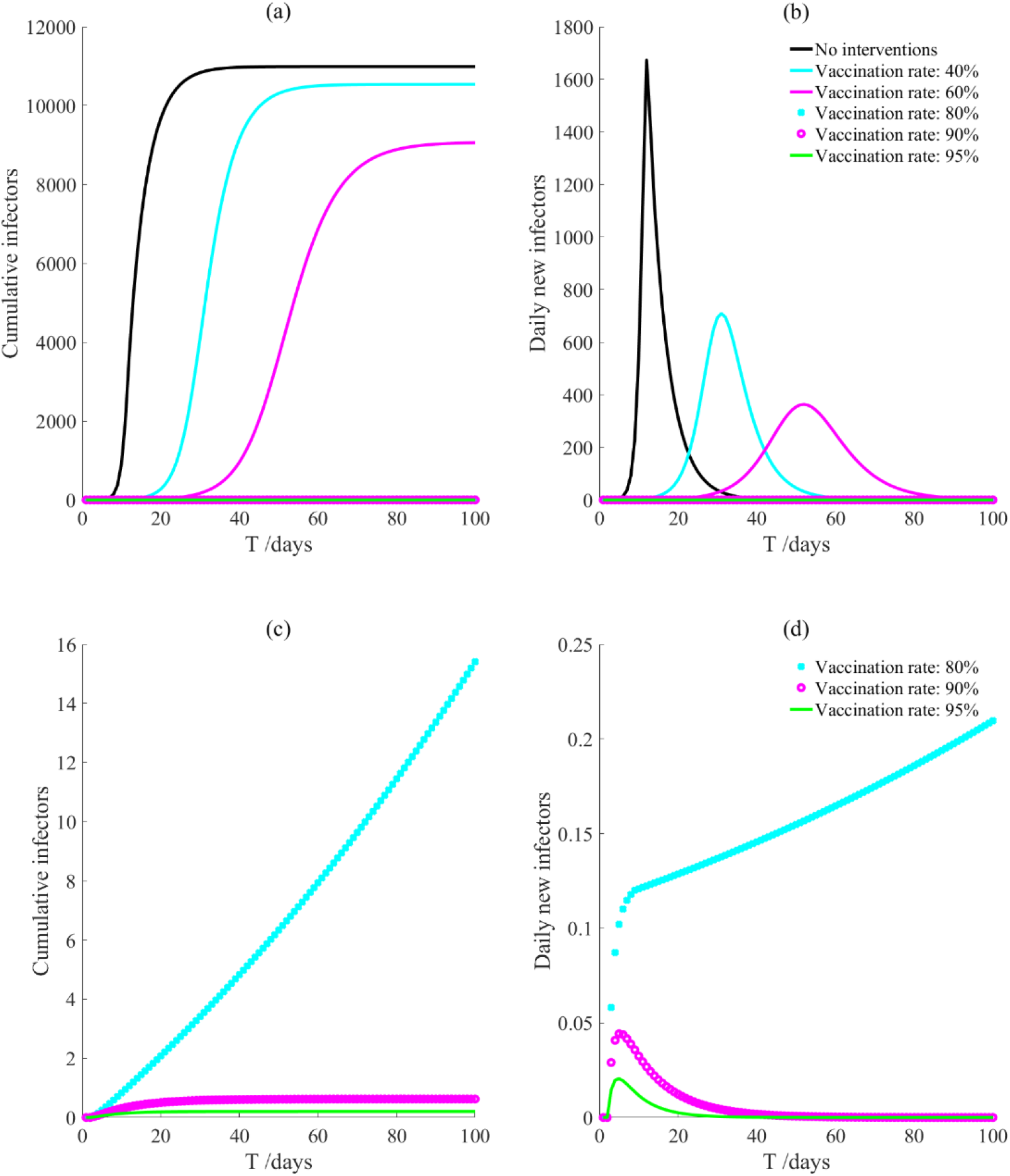
Vaccinated but not quarantined. Assume pre-flight screening can identify 20% of asymptomatic infectors.

Compared with **Fig. 4** and **Fig. 5, Fig. 6** and **Fig. 7, Fig. 8** and **Fig. 9**, it can be seen that under the intervention measures of “vaccinated but not quarantined”, “vaccination and 7-day compulsory quarantine”, “vaccination and 14-day compulsory quarantine”, the proportion of asymptomatic infected individuals identified by pre-flight screening has little effect on the number of secondarily infected individuals, and the outbreak time was basically the same under different vaccination proportions. However, for the intervention measures in each figure in **Fig. 4** to **Fig. 9**, as the vaccination rate increased, the secondary infectors at peak hour and the peak hour of onset significantly decreased. When the vaccination rate of athletes reached 80%, the number of secondary infectors decreased significantly, controlled within 1, indicating that the immune barrier has been reached.

**Fig. 5.**
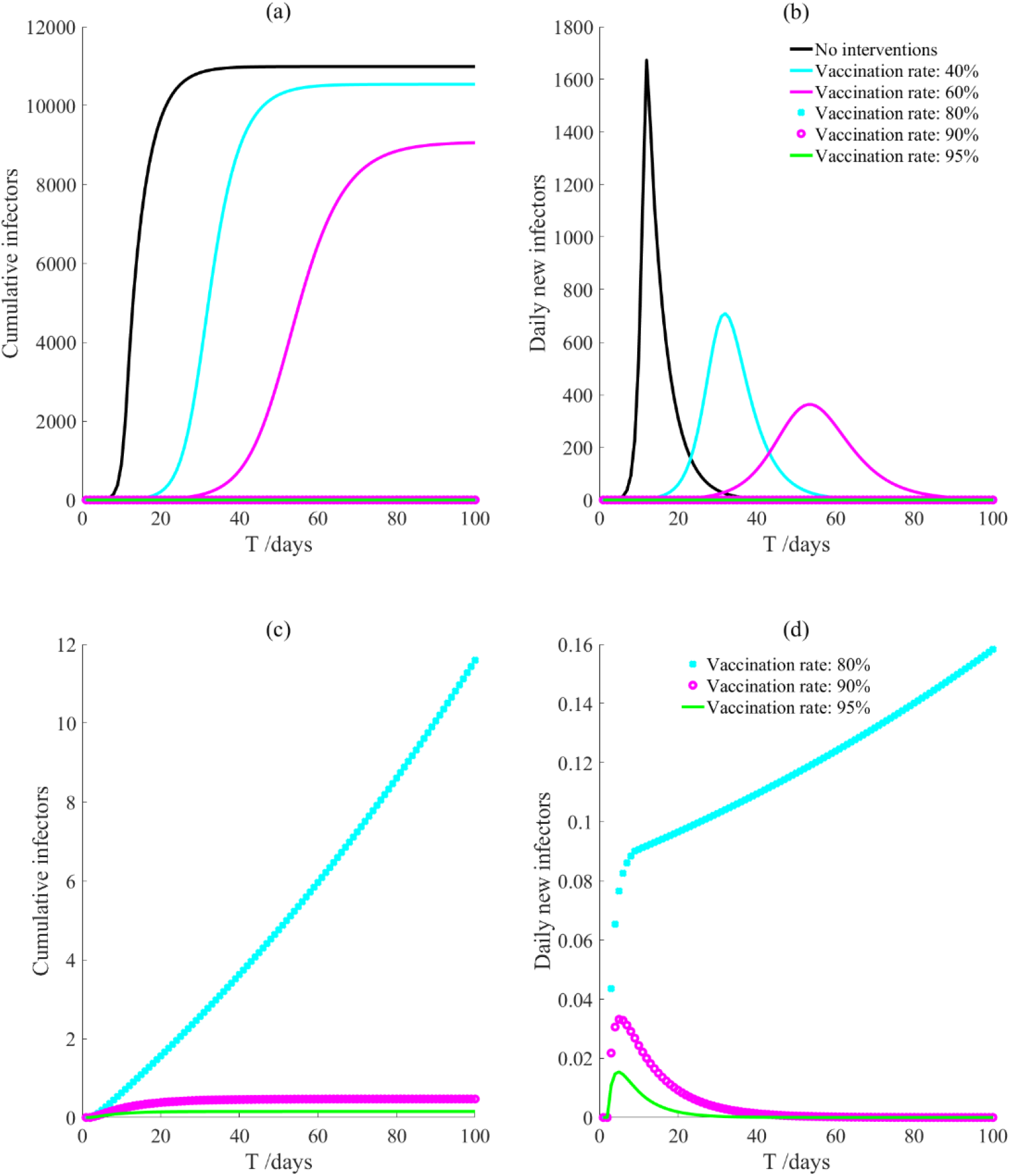
Vaccinated but not quarantined. Assume pre-flight screening can identify 40% of asymptomatic infectors.

**Fig. 6.**
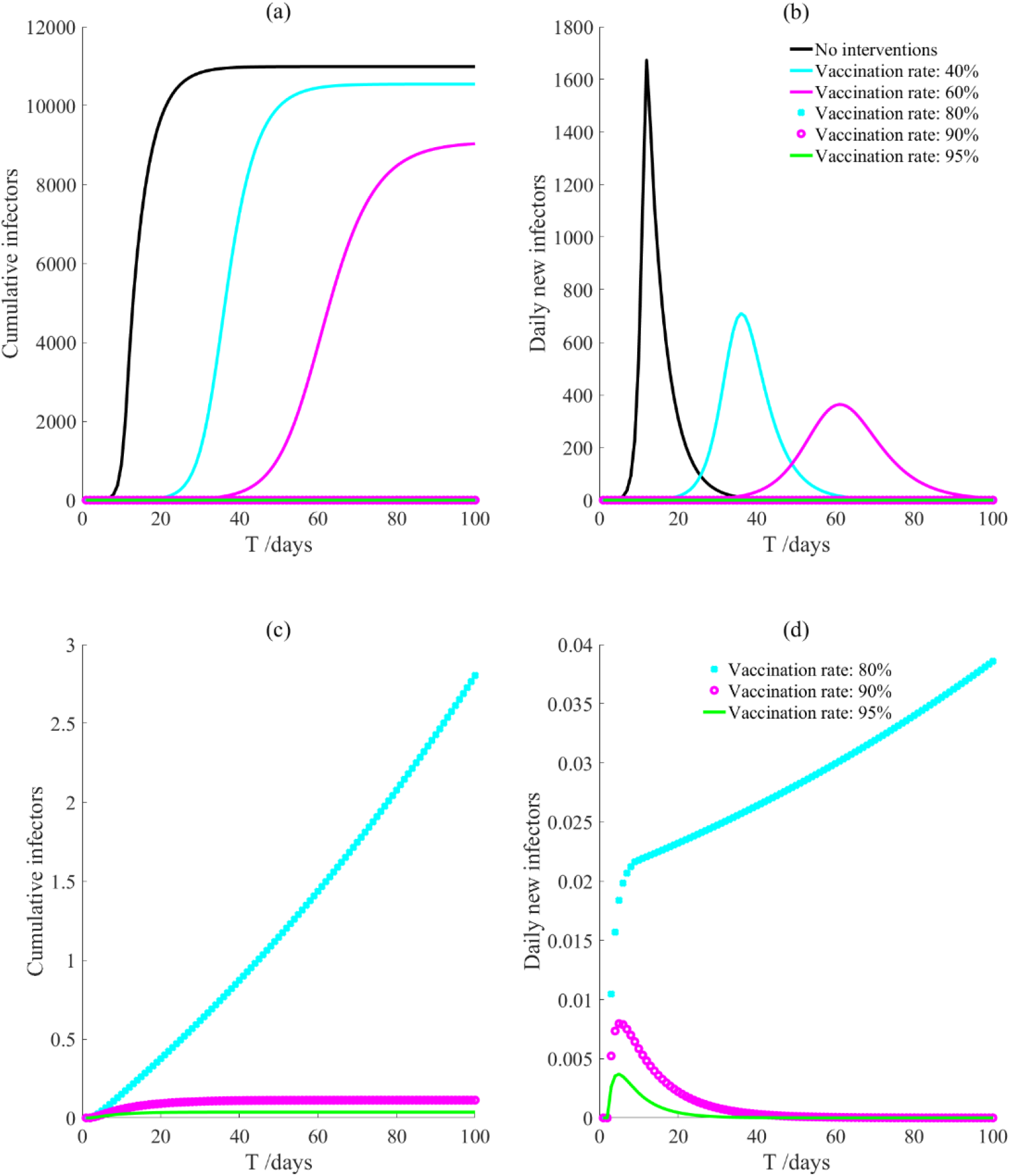
Vaccination and 7-day compulsory quarantine. Assume pre-flight screening can identify 20% of asymptomatic infectors, 7-day compulsory quarantine has about 18% risk of missed detection.

**Fig. 7.**
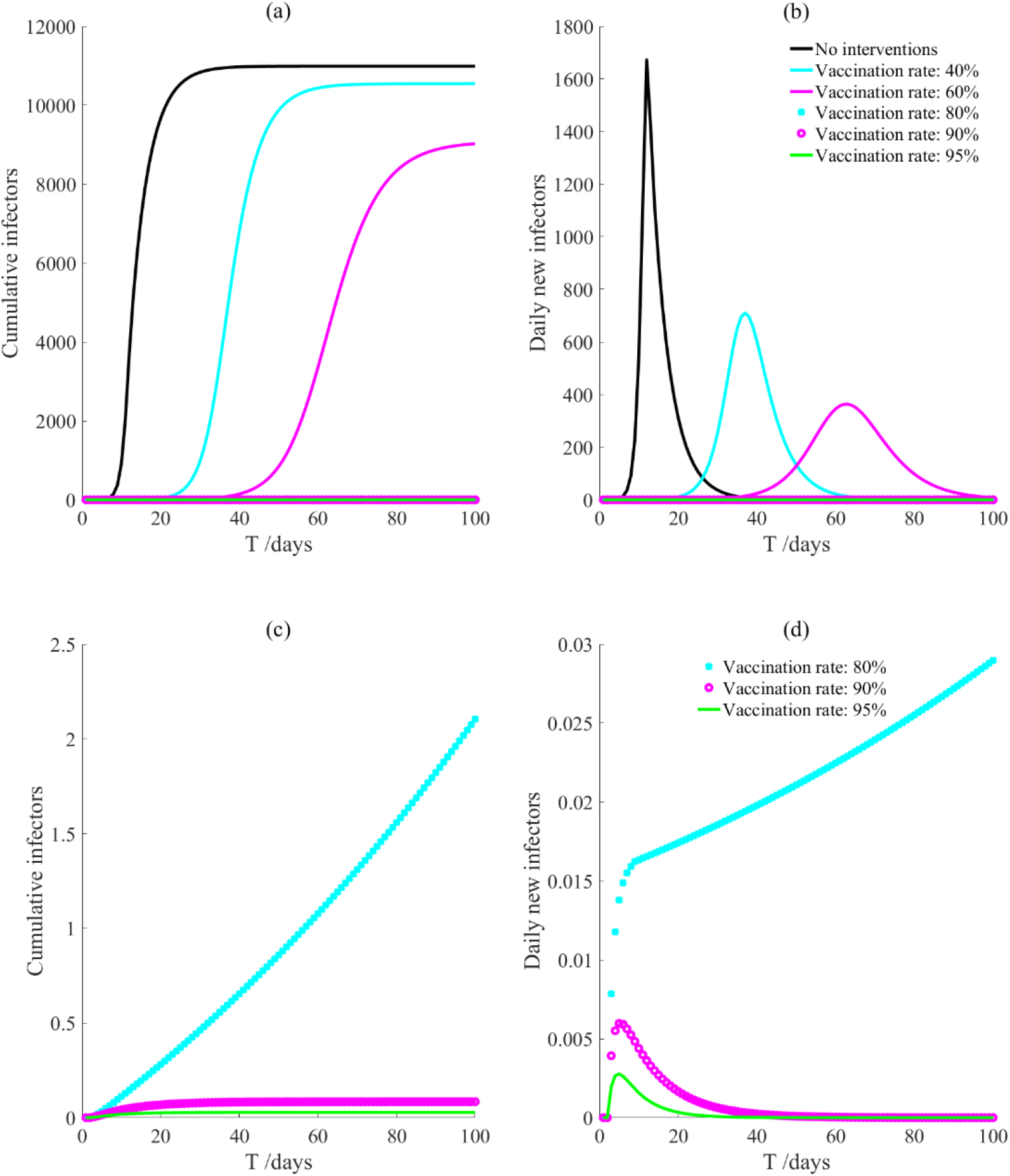
Vaccination and 7-day compulsory quarantine. Assume pre-flight screening can identify 40% of asymptomatic infectors, 7-day compulsory quarantine has about 18% risk of missed detection.

**Fig. 8.**
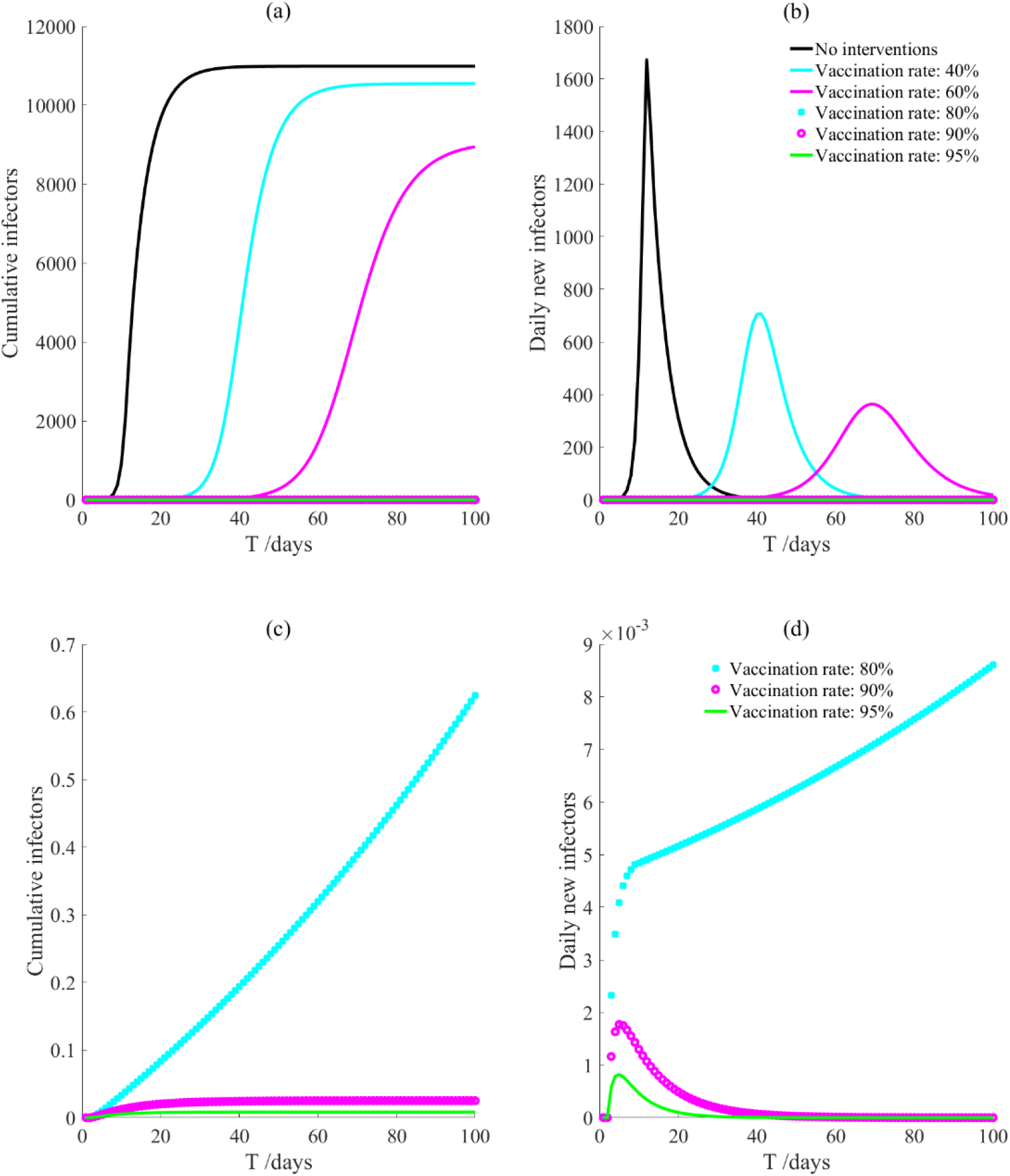
Vaccination and 14-day compulsory quarantine. Assume pre-flight screening can identify 20% of asymptomatic infectors, 14-day compulsory quarantine has about 4% risk of missed detection.

**Fig. 9.**
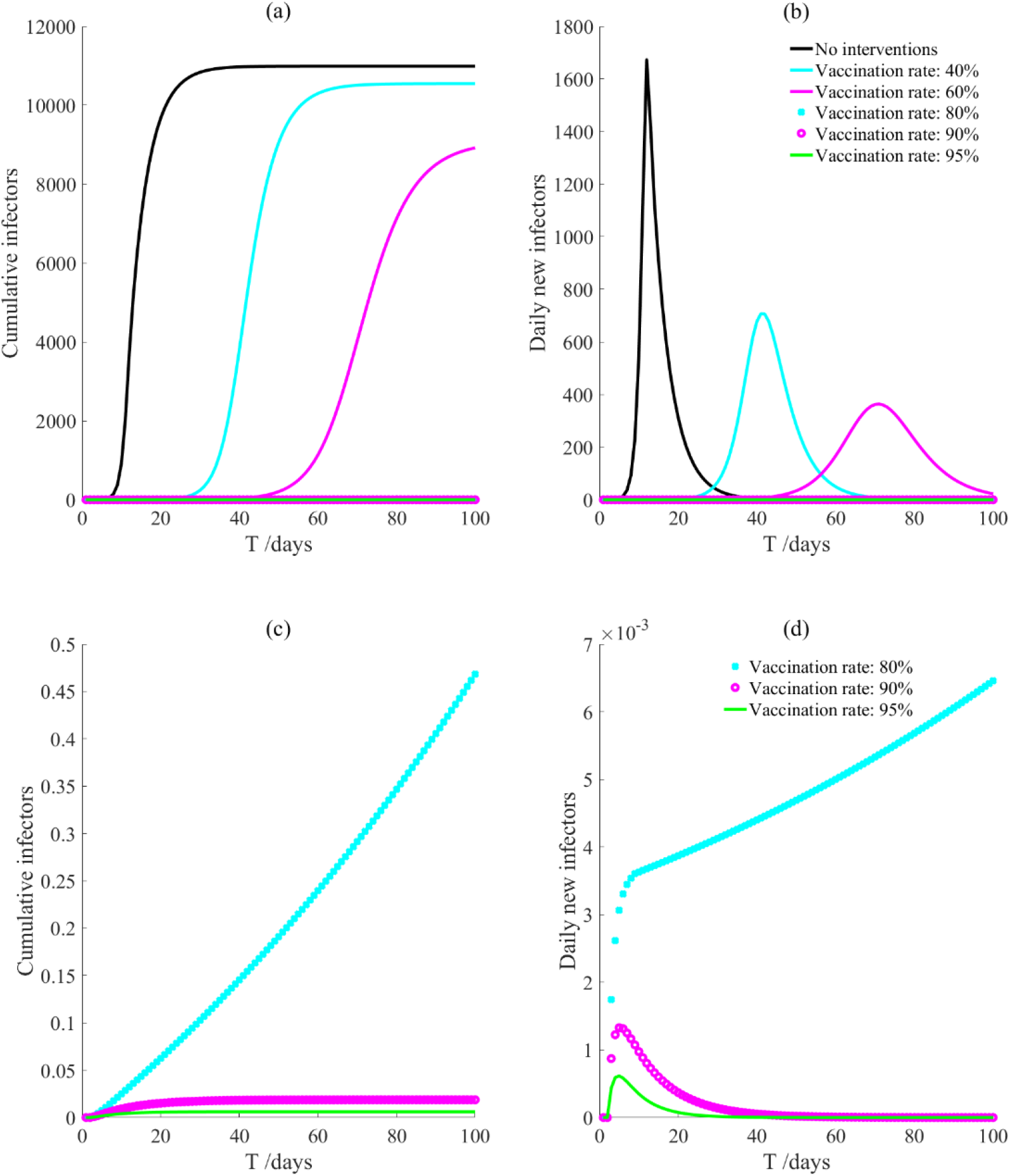
Vaccination and 14-day compulsory quarantine. Assume pre-flight screening can identify 40% of asymptomatic infectors, 14-day compulsory quarantine has about 4% risk of missed detection.

## Discussion

Through the simulation of no intervention measures and various intervention measures, it was found that the prevention and control effect of the measures currently proposed by JOC was not ideal. The total number of secondary infectors can still reach the level of no intervention measure, but the duration increases. However, the prevention and control effect of the intervention measures proposed by us was significantly better than that of no intervention measure and the epidemic response measures proposed by JOC. The number of secondary infectors without intervention measure was 1,673; when the vaccination rate reached 80%, the number of secondary infectors under all intervention measures was less than 1, indicating that the epidemic situation has been effectively controlled, and vaccination can effectively prevent COVID-19. In the face of infectious diseases such as COVID-19 with strong infectivity, in addition to the prevention and control measures listed in this paper, the combination of multiple intervention measures will bring better results. For example, closed-loop management can be adopted during the Olympic Games, sites can be disinfected before the athletes arriving, and the staff can be trained on epidemic prevention and control.

Risk assessment includes risk identification, dose-response relationship, exposure assessment, etc. COVID-19 is mainly transmitted through direct transmission, aerosol transmission and contact transmission. Therefore, this paper starts from the potential risks, namely, intervening in the contact between people, then simulates the potential transmission risks, and finally forms a risk assessment for COVID-19. In this paper, a simulation method of public health intervention prevention and control based on the dynamic model of COVID-19 was proposed, which can be extended to all kinds of large-scale activities or infectious disease prevention and control research. The intervention measures in this paper were based on the epidemic prevention and control measures and ideas proposed by JOC and the IOC. At present, many countries have the ability to produce the COVID-19 vaccine, and IOC will also bear the cost of the vaccine, so it is feasible. Vaccination is not compulsory, so it does not involve human rights intervention, and it is a complex regulation.

The limitation of this study was that there was no specific number of participants from each country, so it was not possible to accurately estimate the initially infected individuals, but we have given the calculation formula. Secondly, this paper assumed that the effective rate of vaccine was 100%, which was not in line with the actual situation, but we solve the problem of vaccine efficiency indirectly through different vaccination rates. But after the IOC published the specific data and obtained the effective rate of the vaccine, we can carry out accurate transmission simulation.

## Conclusions

Based on the dynamic model, this paper simulated different prevention and control measures of Tokyo Olympic Games, comparing the number of secondary infectors under different measures, and found that vaccination had the best prevention and control effect. When the effective rate of vaccine reached 100%, the vaccination rate reached 80%, then the number of secondary infectors can be controlled within 1. Our study will contribute to the formulation of relevant measures by JOC and IOC. In summary, compared with the current public health interventions, the mass vaccination would become a milestone in the control of COVID-19.

## Data Availability

The data we used was public data.

## Statement of ethics

The survey was discussed with the Ethics Committee of West China fourth Hospital, West China School of Public Health, Sichuan University, who reviewed the content. And the ethical approval was waived.

## Conflict of interest

The authors have no conflict of interest to declare.

## Funding

This research work was funded by Sichuan Science and Technology Program (grant numbers 2020YFS0015, 2020YFS0091), Health Commission of Sichuan province (20PJ092), National Natural Science Foundation of China (grant numbers 81602935), Chongqing Science and Technology Program (grant numbers cstc2020jscxcylhX0003), Sichuan University (grant numbers 2018hhf-26, GSSCU2018038) and Liang Shan Zhou Center for Disease Control and Prevention. The funders played no role in the design of the study and collection, analysis, and interpretation of data and in writing the manuscript.

## Author contributions

WHZ consulted the literature, analyzed the data, wrote the programs, and was a major contributor in writing the manuscript. JF wrote part of the manuscript. CL collected the parameters of the model. HMW made comments on the content of the article. YZ collected part of the data. XZ made constructive comments on the manuscript. TZ contributed significantly to analysis and manuscript preparation.

## Acknowledgements

Many thanks are expressed to those who contributed to the prevention and control of COVID-19. The authors also want to thank Hongli Wan for the submission of manuscript.

## Notes

### Competing Interest Statement

The authors have declared no competing interest.

